# FIThe Effects of Curcumin Supplementation on Body Weight, Body Mass Index, and Waist Circumference in Patients with Type 2 Diabetes: A Systematic Review and Meta-Analysis of Randomized Controlled Trials

**DOI:** 10.1101/2024.10.08.24315114

**Authors:** Noorbakhsh Alivand, Soleyman Alivand, Seyed jalil masoumi, Sahar foshati, Ebrahim Abbasi

**Author notes:** Correspondence should be sent to E. Abbasi at or.

## Abstract

The study investigates the effects of curcumin on obesity and its related factors in diabetic patients, given that obesity can lead to insulin resistance and complicate diabetes management. An analysis of nine randomized controlled trials (RCTs) involving 699 participants was conducted to evaluate curcumin’s impact on body weight (BW), body mass index (BMI), and waist circumference (WC). The results indicated a significant reduction in BW (WMD: −1.65 kg), BMI (WMD: −0.69 kg/m2), and WC (WMD: - 0.93 cm) with curcumin consumption compared to controls. Higher doses of curcumin (>1000 mg/day) were notably more effective in reducing BW, BMI, and WC than lower doses. The study concludes that curcumin supplementation effectively reduces BW, BMI, and WC in diabetic patients, especially at higher doses, and suggests the need for further studies to explore these benefits.

## Introduction

Diabetes mellitus is a group of metabolic diseases involving hyperglycemia due to defects in insulin secretion, action, or both or insulin resistance in skeletal muscle, liver, and adipose tissues, with a failure of B-cell compensation and a relative insulin deficiency[1].

Recently, the incidence of diabetes has increased, and it is expected that the prevalence of type 2 diabetes among adults will increase from 285 million patients in 2010 to 438 million patients in 2030. More than 60% of diabetic patients live in Asia, and there are more than 4 million type 2 diabetic patients in Iran[2]. Besides, almost 90% of T2DM patients are overweight or obese[3].

Obesity, in particular in the form of abdominal adiposity, is known to be associated with increased morbidity and mortality[4]. Diabetes is an obesity-associated disease linked to insulin resistance and hyperglycemia[5]. Recent research has shown that obesity affects more than 600 million people worldwide[6]. Additionally, if scientists do not take immediate action, many people will experience obesity-related disorders, such as type **2** diabetes[2]. Obesity and overweight are two major risk factors for diabetes and its complications. Investigations indicate that almost 90% of diabetic patients are overweight. Obesity can affect the metabolism in the body by causing insulin resistance and reducing insulin secretion[7]. Therefore, the management of obesity has been strongly recommended to control diabetes and its complications[3].

Various courses of action, including surgical approaches, several drugs, and lifestyle interventions, have been recommended for weight management; however, the effectiveness of these approaches is under discussion[8]. Weight reduction resulted in a greater control of T2DM[3]. Lifestyle changes, i.e., supplementation and exercise, are among the complementary treatment approaches in patients suffering from T2DM. (Diabetes Mellitus 2Type) Curcumin is one of the supplements with positive effects noticed in weight control[9, 10].

Curcumin is the most important component of turmeric (Curcuma longa) and is yellow. It is commonly used as a food additive in Asian countries[11]. It is also deemed responsible for many medical properties of turmeric. Curcumin has low toxicity and performs various pharmacological functions, including antioxidant, anti-inflammatory, anti-microbial, and anti-carcinogenic effects[6, 12, 13]. In some clinical trials, the desirable effects of curcumin have been shown to be a therapeutic agent for the treatment of diabetes and complications of high blood glucose[6].

Moreover, several studies reported conflicting effects of curcumin consumption on body weight and composition. Some clinical trials proposed a significant effect of curcumin intake on indicators of body composition[6, 14–16]. On the contrary, others reported no significant effect of curcumin intake on body weight (BW) or body mass index (BMI)[3, 17, 18]. However, despite various studies conducted on the effect of curcumin intake on body composition in patients with type 2 diabetes, no prior study has examined BW, BMI, and WC solely in diabetic patients. Thus, the current study was done to perform a comprehensive systematic review and meta-analysis of published randomized controlled trials (RCTs) to independently evaluate the effect of curcumin supplementation on body weight (BW), body mass index (BMI), and waist circumference (WC) in patients with type 2 diabetes.

## Materials and Methods

The design of this systematic review and meta-analysis has been prepared according to the guidelines of the **2020** Preferred Reporting Items for Systematic Reviews and Meta-Analyses (PRISMA, **2020**) statement[19]. Furthermore, reports of included and excluded primary studies in the different stages of the current systematic review have been presented using the PRISMA flow chart.

### Search strategy

Our meta-analysis was designed according to the guidelines of the PRISMA statement[19]. PubMed, Scopus, and ISI, Web of Sciences databases were searched for English-language reports of relevant RCTs published till January 2024 for related published articles evaluating the effects of curcumin supplementation on body weight, body mass index, and waist circumference. The following MeSH terms and related keywords were searched: “Curcumin “OR “Curcuminoid” OR “Curcuma” OR “Turmeric” OR “Tumeric” OR “Curcuma longa “OR “C. longa” OR “Curcumin longa L.” Details of our search strategy are shown in Appendix S1. The search was not limited to the English language or human subjects. Moreover, we hand-searched the reference list of included articles, related reviews, and meta-analyses. PubMed’s ‘My NCBI’ (National Centre for Biotechnology Information) email alert service was also created to identify new articles that may be published after our search.

### Study selection

Two investigators (N.AL and S.A) reviewed the titles and abstracts of all identified studies to ascertain whether these studies are eligible for this meta-analysis based on our inclusion criteria. Discrepancies were resolved by discussion with S.F. Studies were chosen for analysis according to the following inclusion criteria: 1- the study was a placebo-controlled trial with either a parallel or crossover design; 2- conducted in patients with type 2 diabetes; 3- the effects of curcumin on body weight, body mass index and waist circumference could be extracted from the article (adequate information on body indices with standard deviations (SDs), standard errors (SEs), or 95% CI, at baseline and the end of follow-up in intervention and control group); 4- the studies with an appropriate controlled design, i.e., the only difference between the control and intervention groups was curcumin; and 5- having an intervention duration of at least four weeks. Studies were excluded if 1- we could not extract the net effect of curcumin; 2- curcumin supplementation duration was < 4 weeks; and 3- non-RCT studies. Furthermore, studies with animal, case-control, cross-sectional, or cohort designs, lacking necessary data for extraction, and trials without appropriate controlled design, i.e., those articles using curcumin in combination with other supplements, were excluded from this meta-analysis.

### 2.3. Data extraction

The following information was extracted from each of the eligible RCTs by two independent authors (N.AL and F.S): first author’s name, study location, year of publication, the sample size in each group, type, and dose of intervention and placebo, duration of the intervention, study design (crossover or parallel), patient’s status and other information including age, gender, and baseline BMI. We also extracted the mean and SD values of body parameters at the study baseline and end of the study. If the data were reported at multiple measurements, only the values at the end of trials were used. In the studies that used only one specific curcumin dose, the analysis included the dose.

### 2.4. Quality assessment

The risk of bias tool was applied using criteria outlined in the Cochrane Handbook for Systematic Reviews of Interventions to assess methodological quality[20]. Two independent reviewers (N.AL and FS) investigated the following criteria for each included study: random sequence generation, allocation concealment, blinding of participants and personnel, blinding of outcome assessment, incomplete outcome data, selective outcome reporting, and other potential sources of biases. Any disagreement in the quality assessment process was resolved through discussion and consultation with a third researcher (S.F.).

### Quantitative data synthesis and statistical analysis

We evaluated the influence of curcumin supplementation on change in the following outcomes: (I) body weight (kg), (II) BMI (kg /m2), and (III) WC (cm). Effect sizes for the meta-analysis were defined as weighted mean difference (WMD; value at the end of the trial minus the value at baseline) and 95% CI[21]. In the event of no reported SD of the mean difference, it was calculated as follows:

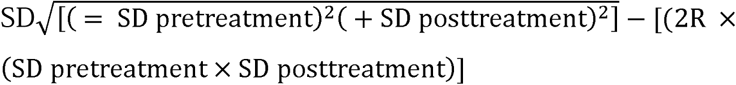

In which a correlation coefficient of 0.5 was assumed as this R-value is a conservative estimate between 0 and 1[22]. When SE was reported instead of SD, we converted SE to SD for analyses: 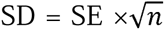, where n is the number of participants in each group. If interested outcomes were reported as median and range, all mean and SD values were estimated using the method described by Hozo et al[23]. Plot digitizer software was used to extract data only when the outcome variable was presented in graphic form. Pre-defined subgroup analyses were performed on supplementation duration and supplement dosage.

Random-effects meta-regression analysis was performed using an unrestricted maximum likelihood method to explore the association between changes in body weight, body mass index, duration, and dose of curcumin supplementation. Statistical heterogeneity between studies was also evaluated using Cochran’s Q-test (significance set at p < 0.1) and I^2^ (≥ 50% assumed to indicate substantial heterogeneity among studies). In the presence of heterogeneity, the pooled effect size was calculated using a random-effects model; otherwise, we applied a fixed-effects model. Sensitivity analysis was used to explore the extent to which inferences might depend on a particular study using the leave-one-out method (i.e., removing a single trial at a time and repeating the analyses[24]. Publication bias was assessed using funnel plots, Begg’s rank correlation, and Egger’s weighted regression tests. In the event of publication bias, the Duval and Tweedie ‘trim and fill ‘and ‘fail-safe N ‘methods were utilized[25]. All statistical analyses were performed using Comprehensive Meta-Analysis (CMA) V3 software with a significance level of p < 0.05.

## Results

### Selection of trials

The explicit research screening process is shown in Figure 1. The preliminary and additional database retrieval yielded 2925 records, with 459 articles excluded due to duplicated records and 274 articles due to review records. A total of 2110 items were eliminated after reading the titles and abstracts following the inclusion and exclusion criteria. After reading the full texts of 82 studies, seventy-three studies were excluded for the following reasons: duplicate report (n = 3), not randomized placebo-controlled studies (n = 5), not providing enough information (n = 38), animal study (n = 3), vitro study (n = 3), review study (n = 2), no anthropometric measurements performed (n = 17) and use of curcumin in combination with other components without an appropriate control group (n = 2). Nine eligible RCTs were included in the meta-analyses[1–3, 6, 10, 13, 26–28]. Of these, six articles reported the effects of curcumin on BW, eight articles on BMI, and three on WC.

**Fig 1.**
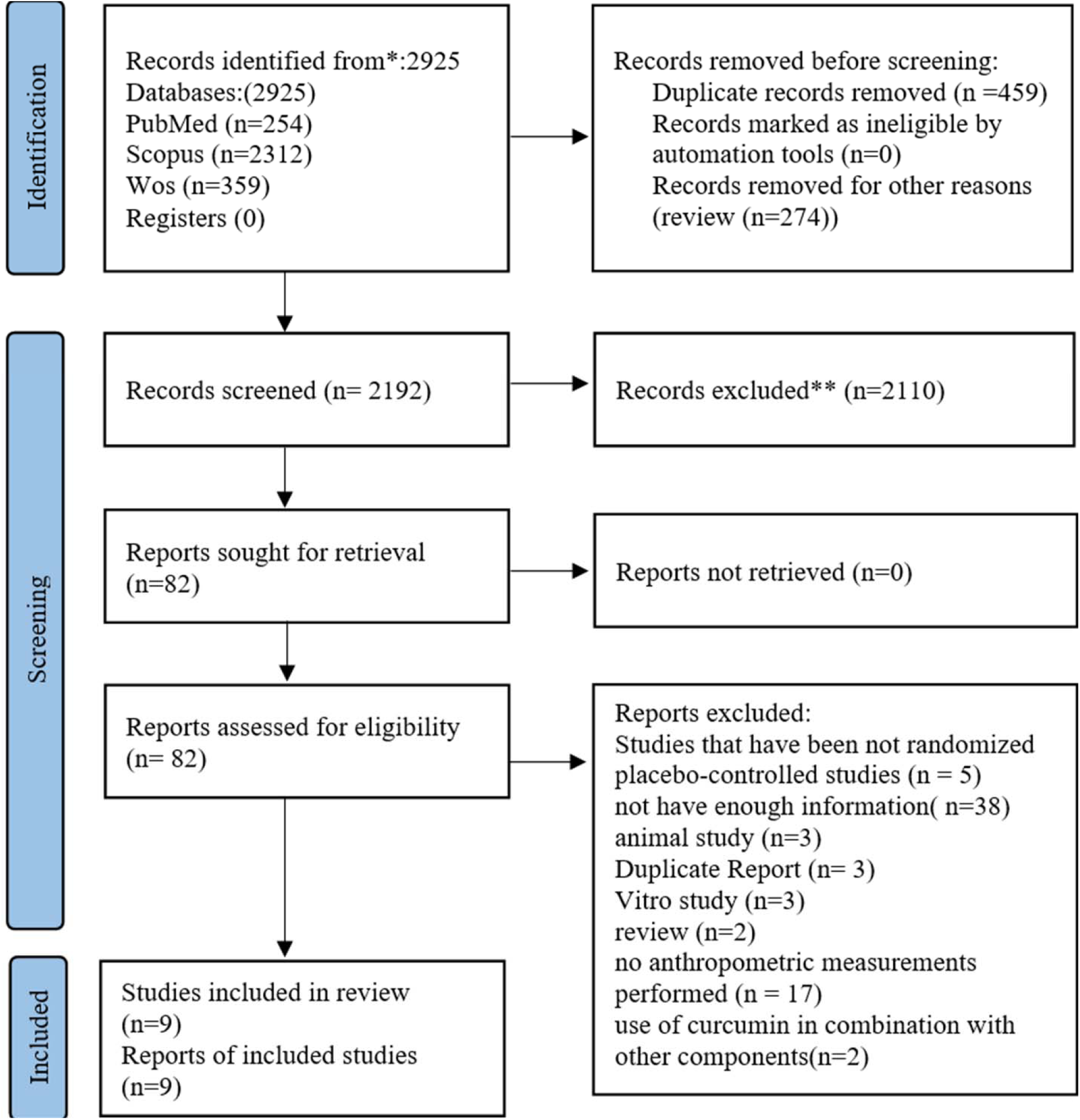
The review process based on PRISMA flow chart

### Characteristics of the included studies

Study characteristics of the 9 eligible articles are presented in Table 1. Data were pooled from these studies, and the sample size ranged from 21 to 213. Overall, 1,088 participants were randomly assigned in these trials, and 699 participants (64.24%) completed the studies. The mean age of the participants ranged from 43 to 62.5 years. One of the nine trials was performed exclusively on women [10]. The eight remaining trials included both sexes. Eligible studies were published between 2014 and 2021, most of which were conducted in Iran. The remaining studies were conducted in Thailand and Brazil. Seven treatment arms used curcumin for the intervention. Cooked rice, Polysorbate capsules, placebo capsules, maltodextrin, placebo capsules containing starch, and placebo + 10 mg piperine were used for the control groups. Two treatment arms supplemented turmeric for the intervention, while the control group was provided with cornstarch flour. A wide range of curcumin supplementation doses was utilized in the study designs: curcumin capsules of 80 to 1500 mg/d and turmeric capsules of 2100 mg per day. Supplementation duration varied from 8 to 28 weeks. Participant characteristics were also similar between studies, focusing on specific patient populations, including type 2 diabetic patients (Table 2).

**Table 1.** Main characteristics of the included studies.

| First author<br>(publication<br>year) | Country | Sample size<br>(M/F,<br>Intervention/<br>Control) | Target<br>population | Mean<br>age<br>(yr) | BMI<br>(kg/m<sup>2</sup>) | Study design<br>(randomization,<br>blinding) | Intervention<br>group | Control<br>group | Dose<br>(mg/d) | Duration<br>(w) | Results |
| --- | --- | --- | --- | --- | --- | --- | --- | --- | --- | --- | --- |
| Hamid Reza Rahimi (2015) | Iran | 70 (31/39,<br>35/35) | T2DM | 56.3 | - | Parallel (yes,<br>double) | Curcumin | placebo | 80 | 12 | ↓ BMI |
| Homa Hodaiei (2019) | Iran | 44 (22/22,<br>21/23) | T2DM | **58** | 18.5-35 | Parallel (yes,<br>double) | Curcumin<br>capsules | cooked rice | 1500 | 10 | ↓ BW, ↓<br>BMI,<br>↓ WC, |
| Joana Furtado de Figueiredo Neta, (2021) | Brazil | 61 (14/47,<br>33/28) | T2DM | 62.5 | - | Parallel (yes,<br>double) | curcumin<br>capsules plus<br>5mg piperine | Placebo<br>capsules | 500 | 17 | ↓ BMI, |
| Mahsa Ahmadi Darmian (2021) | Iran | 21 (0/21,<br>11/10) | T2DM | 44.3 | 25-30 | Parallel (yes,<br>single) | turmeric<br>capsules | Corn starch<br>flour capsules | 2100 | 8 | ↓ BW, ↓<br>BMI, |
| Sara Asadia (2019) | Iran | 80 (10/70,<br>40/40) | T2DM | 53.3 | 25-39.9 | Parallel (yes,<br>double) | curcumin<br>capsules | Polysorbate<br>capsules | 80 | 8 | ↓ BW, ↓<br>BMI,<br>↓ WC |
| Shirin Ebrahimkhani (2020) | Iran | 35 (13/22,<br>16/19) | T2DM | 56.1 | - | Parallel (yes,<br>double) | curcumin | maltodextrin | 500 | 12 | ↔, BW,<br>↔, BMI, |
| Somlak Chuengsamarn (2014) | Thailand | 213 (/97,116<br>107/106) | T2DM | 59.1 | - | Parallel (yes,<br>double) | curcumin<br>capsules | placebo<br>capsule<br>contains starch | 1500 | 25 | ↓ WC, |
| Yunes Panahi (2017) | Iran | 100 (51/49,<br>50/50) | T2DM | 43 | - | Parallel (yes,<br>double) | curcumin plus<br>10 mg piperine | PLASEBO+<br>10MG<br>PIPERINE | 1000 | 12 | ↓ BW, ↓<br>BMI, |
| Zohreh Adab (2018) | Iran | 75 (36/39,<br>36/39) | T2DM | 54.7 | - | parallel (yes,<br>double) | turmeric<br>supplement | corn starch<br>flour | 2100 | 8 | ↓ BW, ↓<br>BMI, |

Symbols: ↓, significant decrease; ↑, significant increase; ↔, no significant difference.
Abbreviations: BW; body wight, BMI, body mass index; WC, waist circumference

**Table 2.**
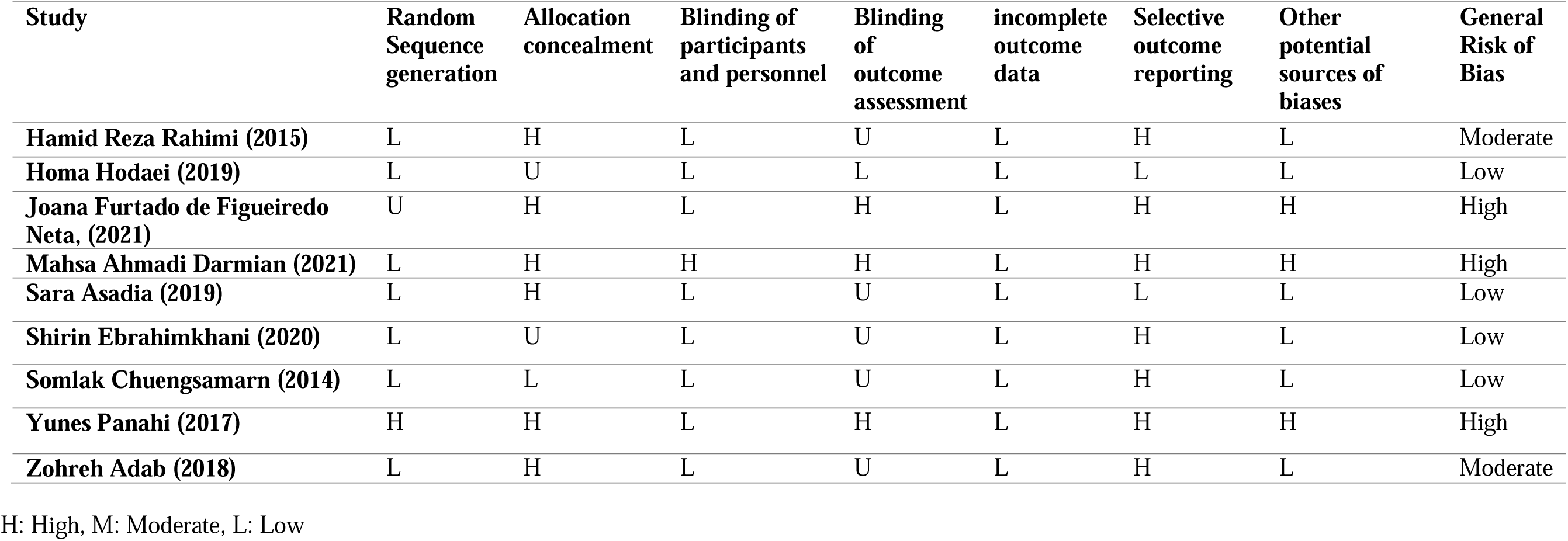
*Quality assessment of clinical trials (according to Cochrane guideline)* The effects of curcumin supplementation on body wight, body mass index and waist circumference in patients with type 2 diabetes.

### Risk of bias

We evaluated the quality of evidence using the Grading of Recommendations, Assessment, Development, and Evaluation (GRADE) approach[29]. GRADE consists of a rating system where the quality of the evidence for each outcome ranges from high to very low. The grade results are shown in Table 3. To sum up, our results indicated a moderate effect of curcumin on body weight, a low effect on BMI, and a high effect on WC.

**Table 3.** Grade profile of Curcumin supplementation on Body weight, BMI and WC.

| Outcomes | Quality assessment |  |  |  |  | Summary of findings |  |  |
| --- | --- | --- | --- | --- | --- | --- | --- | --- |
|  | Risk of bias | Inconsistency | Indirectness | Imprecision | Publication bias | Number of intervention/control | WMD, 95% CI | Quality of evidence |
| ***Body weight*** | NO Serious limitation | Serious limitation | NO Serious limitation | NO Serious limitation | NO Serious limitation | 174/181 | -1.65(-3.04, -0.27) | moderate |
| ***BMI*** | NO Serious limitation | Very Serious limitation | NO Serious limitation | NO Serious limitation | NO Serious limitation | 242/244 | -0.69(-0.99, -0.39) | Low |
| ***WC*** | NO Serious limitation | NO Serious limitation | NO Serious limitation | NO Serious limitation | NO Serious limitation | 168/169 | -0.93(-1.42, -0.44) | High |

## Summary of findings

### The pooled estimate of curcumins on body weight, body mass index, and waist circumference indices

The results of forest plots are illustrated in Figure 2A–C. The body weight, BMI, and WC results were reported using the random-effect model because statistically significant heterogeneity was observed between these studies. Combined results from six studies (174 cases and 181 controls) indicated a significant decrease in body weight after curcumin consumption (WMD: - 1.65 kg, 95% CI: −3.04, −0.27, p = 0.019) with significant heterogenicity between the studies (I^2^ = 78.67%, p = 0.000) (Fig. 2A).

**Figure 2A.**
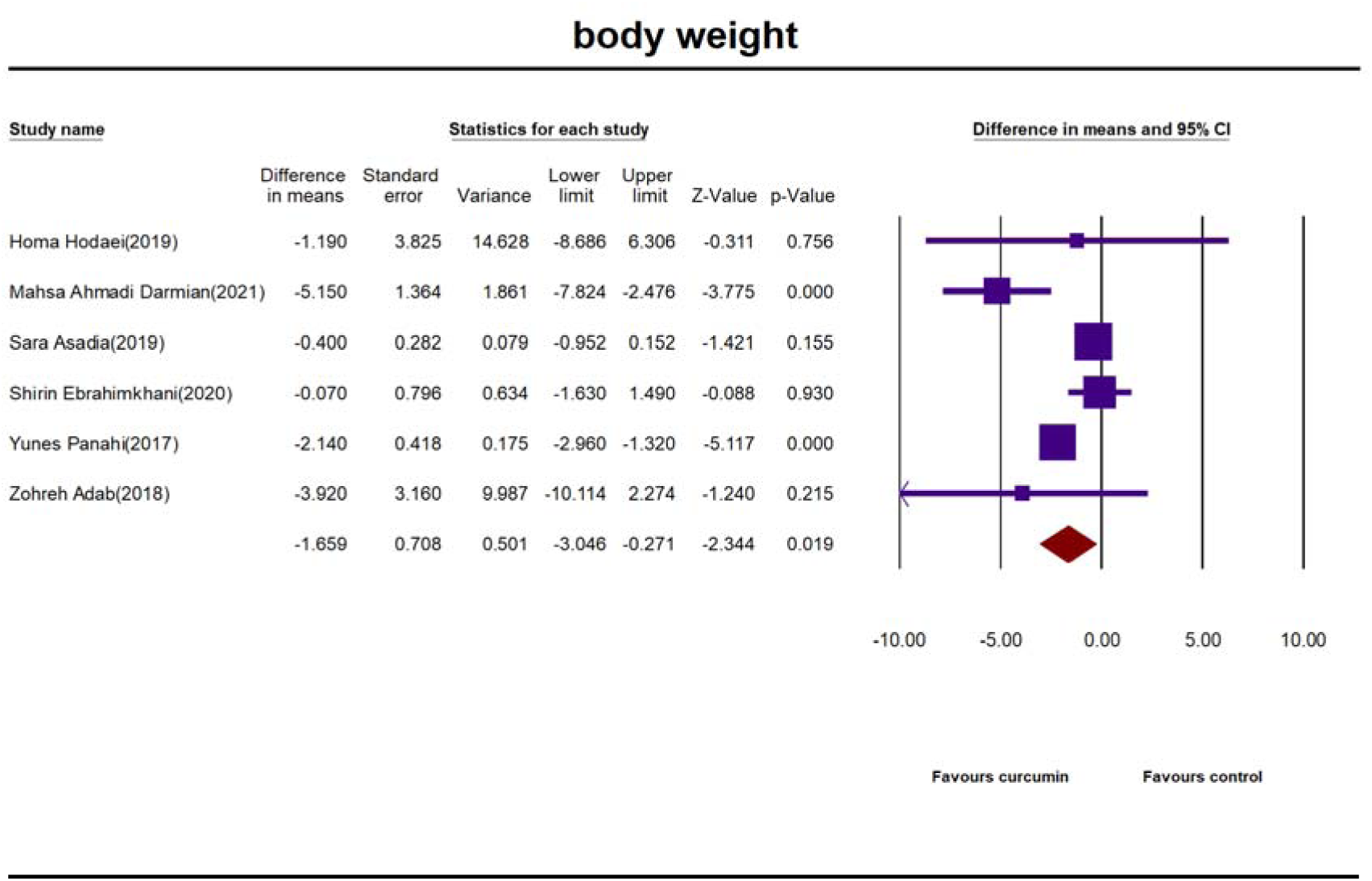
The body weight, BMI, and WC results were reported using the random-effect model.

Pooled results from eight trials (242 cases and 244 controls) showed a significant decrease in BMI in curcumin consumers compared with the control group (WMD: - 0.69 kg m2, 95% CI: −0.99, −0.39, p = 0.000), with severe heterogeneity being detected (I^2^ = 92.48%, p = 0.000) (Fig. 2B).

**Fig 2B.**
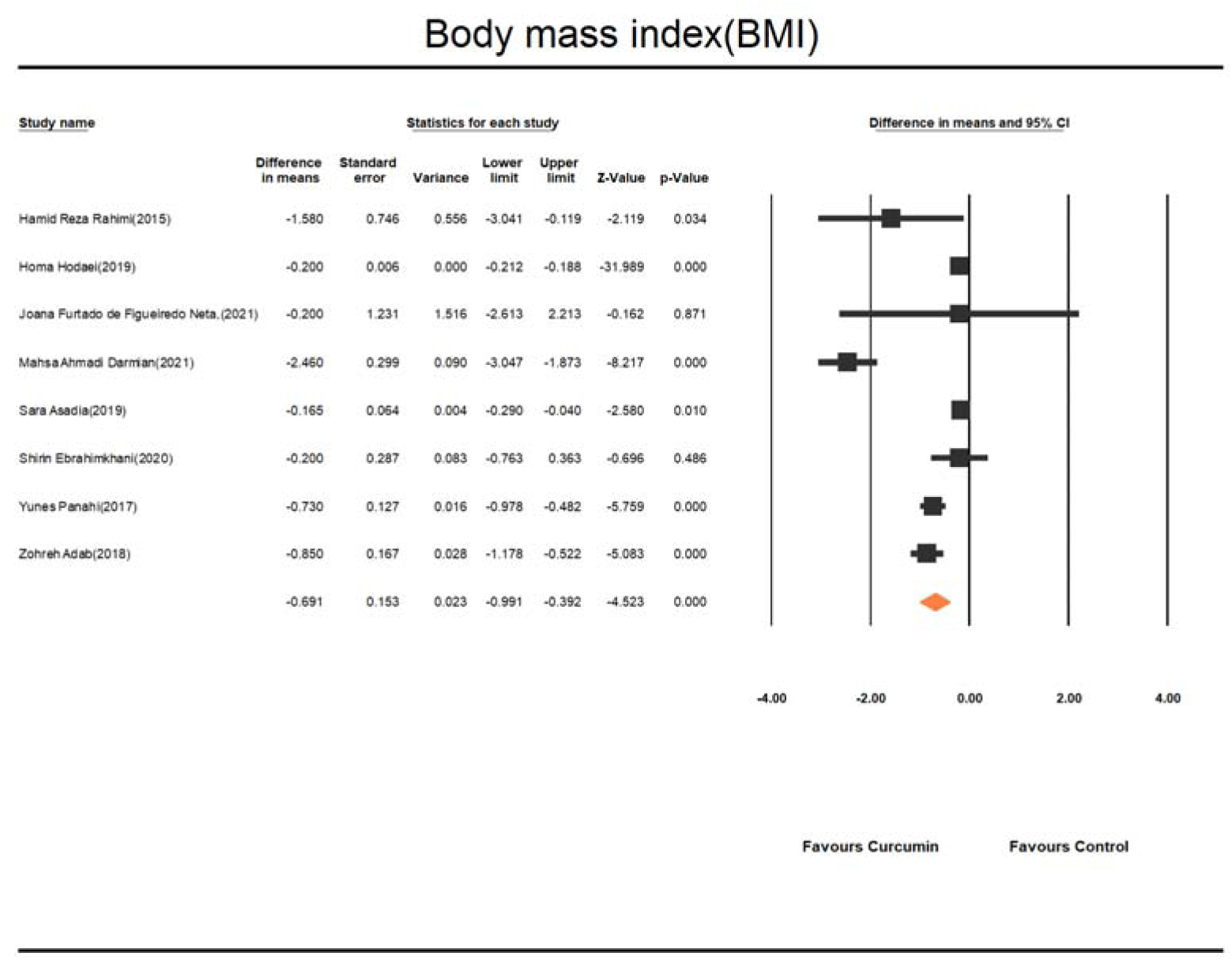
Pooled results from eight trials showed a significant decrease in BMI in curcumin consumer compared with the control group.

Combined results of three studies (168 cases and 169 controls) revealed a significant reduction in WC percent following curcumin consumption (WMD: - 0.93cm, 95% CI: −1.42, - 0.44, p = 0.000). There was no significant heterogenicity between the studies (I^2^ = 25.65%, p = 0.261) (Fig. 2C).

**Fig 2C.**
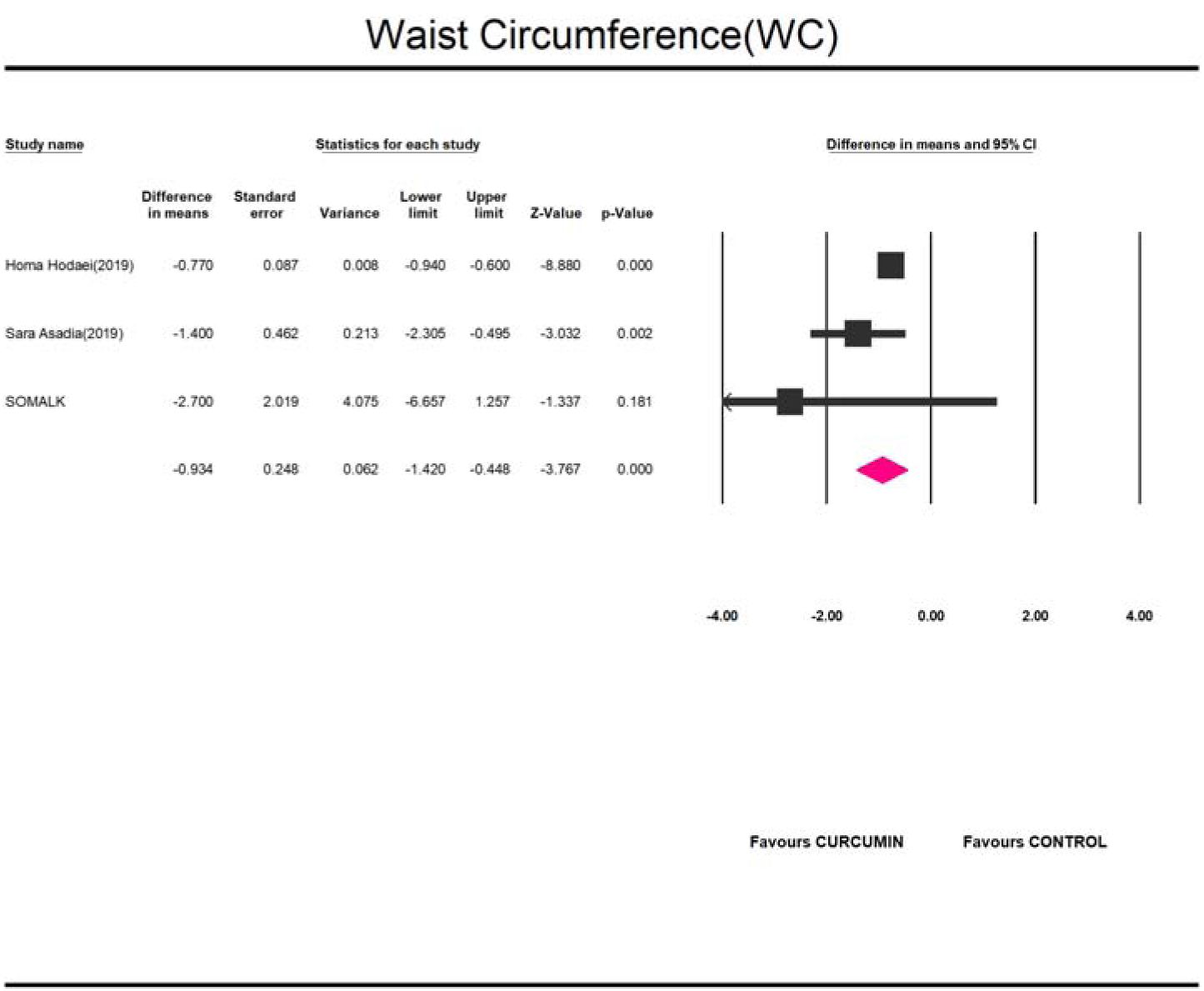
Combined results of three studies revealed a significant reduction in WC percent following curcumin consumption.

### Sensitivity analysis

To observe the impact of each single study on the pooled effect size, we conducted a leave-one-out sensitivity analysis (Figs. 1A-C). The estimated effect size for curcumin’s effect on BMI and WC was robust, suggesting that removing each trial did not change the analysis results. However, by removing studies of Mahsa Ahmadi Darmian [10] (WMD: −1.04 kg, 95% CI: −2.25, 0.16, p = 0.089) and Yunes Panahi [26], (WMD: −1.59 kg, 95% CI: −3.04, 0.23, p = 0.088), the effect on body weight was no longer statistically significant.

### Subgroup analysis

The subgroup analyses were conducted (Table 4) to explore the effect of curcumin dose and duration on the overall impact of curcumin on the estimated effect size. When the studies were stratified according to the dose of curcumin consumed, as a high dose (> 1000mg/day) or a low dose (≤1000 mg/day), significant reductions in body weight were observed in the subsets of studies with > 1000 mg/day of curcumin consumption (WMD: - 4.59 kg, 95% CI: −6.92, −2.25, p = 0.002) but not in those with ≤ 1000 g/day (WMD: - 0.93 kg, 95% CI: −2.24, 0.37, p = 0.164). Also, curcumin significantly reduced BMI in doses > 1000 mg/day (WMD: −0.99, 95% CI: −1.63, −0.35, p < 0.002). This effect has also been seen in doses ≤ 1000 mg/day (WMD: −0.21, 95% CI: −0.49, 0.05, p < 0.011). The reduction in WC was observed both after consuming > 1000 mg/day of curcumin dose (WMD: −0.77 cm, 95% CI: - 0.94, −0.60, p = 0.000) and lower doses (WMD: −1.04 kg, 95% CI −2.30, −0.49, p = 0.002). However, it seems that the reduction in the higher doses was more than the lower dose.s Subgroup analysis was also performed by dividing the duration of curcumin intervention (≥ 10 weeks vs. < 10 weeks). Subgroup analysis indicated that curcumin consumption does not have a significant effect on reducing body weight (WMD: −1.51 kg, 95% CI: −3.09, 0.07, p = 0.06), BMI (WMD: −0.61, 95% CI: −1.03, −0.19, p < 0.004) and WC (WMD: - 2.70, 95% CI: 6.65, 1.25, p = 0.18) in longer-term interventions.

**Table 4.** Results of subgroup analysis of included randomized controlled trials in the meta-analysis of curcumin supplementation and Body wight, BMI and WC - related parameters.

| **Variables** | **Dose (mg/day)** |  | **Duration (weeks)** |  |
| --- | --- | --- | --- | --- |
| **Body wigt (Kg)** | **>1000** | **≤1000** | **≥10 weeks** | **<10 weeks** |
| **Number of Comparisons** | 3 | 3 | 3 | 3 |
| **WMD (95% CI)** | - 4.59<br>( -6.92, -2.25) | - 0.93<br>(-2.24, 0.37) | -1.24<br>(-2.98, 0.49) | -2.83<br>(-6.68, 1.00) |
| **p value** | 0.002 | 0.164 | 0.161 | 0.148 |
| **I<sup>2</sup> (%)** | 0.605 | 84.63 | 62.38 | 84.28 |
| **P heterogeneity** | 0.000 | 0.001 | 0.070 | 0.002 |
| **BMI (kg/m2)** | **>1000** | **≤1000** | **≥10 weeks** | **<10 weeks** |
| **Number of Comparisons** | 4 | 4 | 4 | 4 |
| **WMD (95% CI)** | -0.99<br>( -1.63, -0.35) | -0.21<br>(-0.49, 0.05) | -0.61<br>(-1.03, -0.19) | -0.73<br>(-1.10, -0.36) |
| **p value** | <0.002 | <0.011 | <0.004 | 0.000 |
| **I<sup>2</sup> (%)** | 96.64 | 16.27 | 33.76 | 95.85 |
| **P heterogeneity** | 0.000 | 0.310 | 0.210 | 0.000 |
| **WC (cm)** | **>1000** | **≤1000** | **≥10 weeks** | **<10 weeks** |
| **Number of Comparisons** | 2 | 1 | 1 | 2 |
| **WMD (95% CI)** | -0.77<br>( - 0.94, -0.60) | -1.04<br>(-2.30, -0.49) | -2.07<br>(-6.65, -1.25) | -0.92<br>(-1.45, -0.39) |
| **p value** | 0.000 | 0.002 | 0.181 | 0.001 |
| **I<sup>2</sup> (%)** | **0.000** | **0.000** | 0.000 | 44.37 |
| **P heterogeneity** | 0.340 | 1.000 | 1.000 | 0.180 |

**Table 5.**
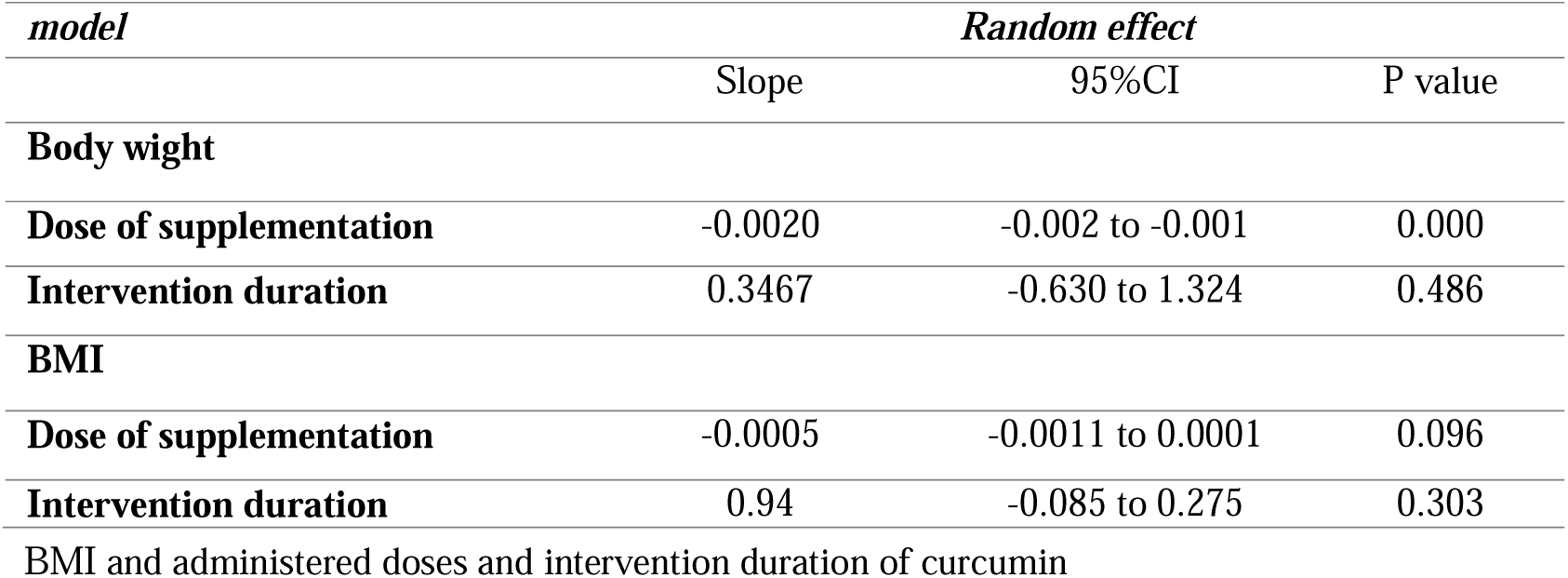
Meta-regression between changes in Body Weight and was found with changes in Body wight and BMI (Fig. 5 panel A-B and Table 3).

| *model* | *Random effect* |  |  |
| --- | --- | --- | --- |
|  | Slope | 95%CI | P value |
| **Body wight** |  |  |  |
| **Dose of supplementation** | -0.0020 | -0.002 to -0.001 | 0.000 |
| **Intervention duration** | 0.3467 | -0.630 to 1.324 | 0.486 |
| **BMI** |  |  |  |
| **Dose of supplementation** | -0.0005 | -0.0011 to 0.0001 | 0.096 |
| **Intervention duration** | 0.94 | -0.085 to 0.275 | 0.303 |
| BMI and administered doses and intervention duration of curcumin |  |  |  |

**Table 6.**
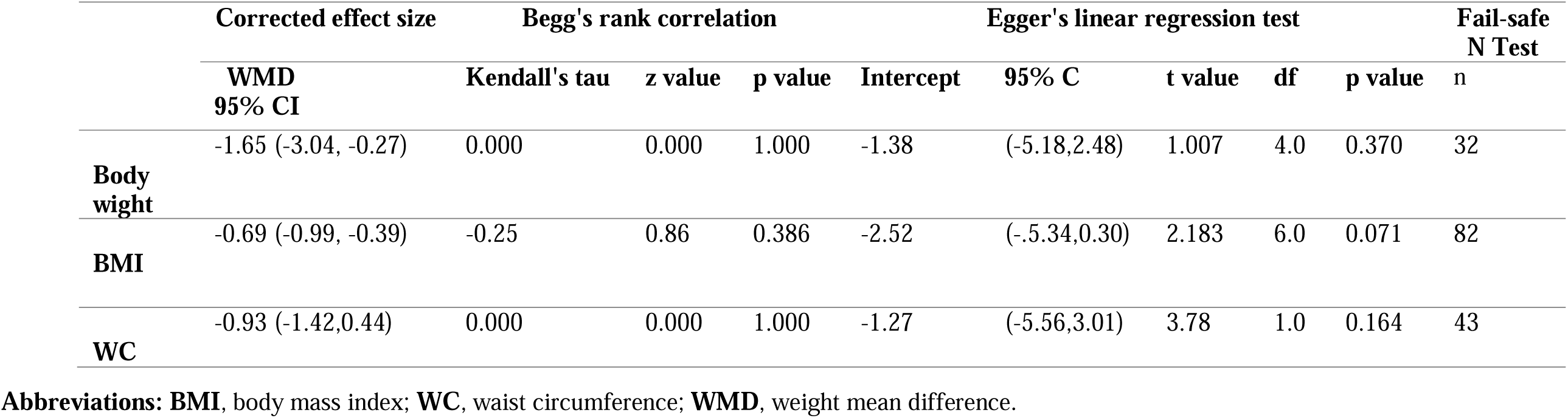
Assessment of publication bias in the impact of curcumin on Body wight, Body mass index and Waist circumference in patients with type 2 diabetes.

### Meta-regression

A significant association was found between changes in body weight and the dose of supplementation, but not with BMI (Fig. S2 panel A-B) and Table 3). Considering the duration of supplementation with curcumin, no significant association was found.

### Publication bias

Based on Egger’s regression test, there was no evidence of publication bias for studies examining the effect of curcumin on body weight (p = 0.370), BMI (p = 0.071), and WC (p = 0.164). Furthermore, based on Begg’s rank correlation tests, there was no significant publication bias for studies on body weight (p = 1.000), BMI (p = 0.386), and WC (p = 1.000). Funnel plots of the impact of curcumin on body weight, BMI, and WC are illustrated in (Fig. S3A-C). Additionally, 32,827 and 43 studies would be required to bring the effect size of body weight, BMI, and WC, respectively, to a non-significant (p > 0.05) value in the analysis of the “fail-safe N” test.

## Discussion

Limited systematic reviews and meta-analyses have been conducted on the effects of curcumin supplementation on body weight, body mass index, and waist circumference. However, the current systematic review and meta-analysis are the first studies that independently evaluate the effect of curcumin supplementation on BW, WC, and BMI from existing RCTs in diabetic patients. A meta-analysis of data from nine RCTs shows a significant effect of curcumin supplementation on BW, BMI, and WC. However, a significant reduction in BW and BMI was observed, especially at doses higher than 1000 mg/day in some trials.

Data obtained from this systematic review and meta-analysis examined curcumin supplementation’s effects on body composition indices, including in adults. In line with our study, Mousavi et al.’s results showed that curcumin supplementation significantly reduced BW and BMI compared to the control group, and unlike our study, no significant effect of curcumin on WC[12]. The findings of a similar systematic review and meta-analysis done by Akbari and Chanita Unhapipatpong demonstrated that curcumin consumption leads to a significantly reduced BW, BMI, and WC, which aligns with the results of our study. However, the reducing effect of curcumin on BW and BMI in our study is more than in those two studies. The effect of curcumin on waist circumference (WC) in our study was found to be greater than that reported by Akbari et al., likely because the participants in our study[30, 31] had similar health conditions. In addition, a systematic review of clinical trials found that curcumin supplementation may reduce obesity and overweight in adults[32]. However, curcumin supplementation did not change BW and BMI in obese adults compared to placebo in a clinical trial[33]. Also, curcumin did not significantly affect BW in patients with metabolic syndrome[17]. Several meta-analyses and clinical trials were conducted, but no significant effect of curcumin on WC[12, 18, 33–35].

Nevertheless, findings from another clinical trial investigating the effect of supplementation with curcumin (1000mg/day) on patients with non-alcoholic fatty liver disease (NAFLD) for 8 weeks showed a significant reduction in WC compared to the supplementation to placebo[36]. It should be noted that, unlike BW and BMI, WC is considered a good indicator of abdominal and general obesity[37]. Due to few available studies on the effect of curcumin supplementation on WC and their conflicting results on the effect of curcumin supplementation on BW, BMI, and WC, further studies are required to reach a firm conclusion in this area[38].

The subgroup analysis reveals some differences, such as significant reductions in BW observed in the subsets of studies with > 1000 mg/day dose of curcumin consumption (p = 0.002) but not in those with ≤ 1000 g/day (p = 0.164). Also, curcumin had a significant effect in reducing BMI in doses > 1000 mg/day (p < 0.002); this effect has also been seen in doses ≤ 1000 mg/day (p < 0.011). The reduction in WC was observed both after consuming doses > 1000 mg/day of curcumin (p = 0.000) and lower doses (p = 0.002). However, it seems that the reduction in the higher dose was more significant than the lower dose. These findings indicate that higher doses of curcumin have a greater effect on anthropometric indices in diabetic patients.

Curcumin is the most important ingredient of turmeric (curcuma longa) and is yellow. Curcumin is comm only used as a food additive in Asian countries [3[6]. This substance also has therapeutic properties. It is an active ingredient of turmeric that has antioxidant and anti-inflammatory properties. Currently, the US Food and Drug Administration (FDA) has approved curcumin as Generally Recognized as Safe (GRAS)[28, 39], and the European Food Safety Authority (EFSA) has recommended an acceptable dose of 3 mg/kg/d[40]. Several studies have shown that curcumin has an acceptable safety profile with minimal side effects[41, 42]. Curcumin has been shown to have several therapeutic benefits for various diseases, such as metabolic syndrome, type 2 diabetes, hyperlipidemia, and Alzheimer’s disease[1, 3, 27, 43]. However, a major limitation of curcumin as an agent with an oral route of administration is its low bioavailability due to its poor absorption in the gastrointestinal tract, rapid metabolism, and fast elimination from systemic circulation[1, 44].

On the other hand, obesity is a global health problem and a major risk factor for type 2 diabetes. Our meta-analysis shows the beneficial effects of curcumin supplementation in reducing BMI, BW, and waist circumference, confirming the results of the latest studies[45].

The mechanisms by which curcumin affects body weight and BMI are unclear. Nonetheless, previous studies suggest several mechanisms through which curcumin may play a role in weight loss. Curcumin down-regulates Janus Kinase (JNK) enzyme, which has been suggested to have a fundamental role in obesity pathogenesis[1, 46, 47]. Curcumin can also inhibit the11bHSD-1enzyme that activates cortisol, and higher cortisol concentrations in adipocytes induce central obesity[48]. In addition, curcumin promotes fatty acid oxidation and increases adipocyte apoptosis. In addition, curcumin inhibits NF-kB in adipose tissue, resulting in a reduced level of inflammatory cytokines, for example, tumor necrosis factor α(TNFa), IL-1, IL-6, monocyte chemotactic protein 1, and plasminogen activator inhibitor type 1. Curcumin has also been revealed to increase adiponectin levels[6, 13, 27]. It reduces obesity by inhibiting adipocyte differentiation in the early stages through the suppression of the transcription factor Proliferator-Activated Receptor-c (PPAR-c) and by activating adenosine monophosphate-activated protein kinase (AMPK), which in turn enhances lipolysis[3, 26].

Finally, the dysbiosis of the intestinal microbiota is also one of the pathogens of obesity, and curcumin acts as a prebiotic, distributing in the intestines and influencing the composition and diversity of the microbiota. However, the effect of curcumin on obesity via gut microbiota homeostasis is still controversia[26, 49, 50]. This study showed that curcumin supplementation can reduce body weight, BMI, and WC in patients with type 2 diabetes, which can effectively reduce insulin sensitivity in type 2 diabetes patients.

### limitations and strengths

Our systematic review has several limitations. Firstly, most of the included RCTs were performed in Iran. Therefore, differences in daily curcumin intake and lifestyle may be underestimated. Secondly, curcumin may have different effects on males and females because of its different fat content, but there was just one article that focused solely on females and no articles on males. Clinical trials are necessary to assess curcumin’s effects on anthropometric variables separately in males and females. Third, the treatment duration in most articles was short. Only two articles reported a study that followed participants for over 12 weeks. Fourth, the significant heterogeneity among studies shows that the effects of curcumin on BW, BMI, and WC should be further investigated. The fifth, the removal of an individual study or a group of studies, as assessed by sensitivity analysis, did not have a significant effect on BMI and WC results. However, the removal of two studies related to BW had a significant effect on the findings. Finally, the number of studies related to WC was less than the appropriate number to perform the regression test, so the regression test was not performed on this index.

It should be noted that this study was not registered with Prospero. This systematic review has some strength. To the best of our knowledge, this study is the first meta-analysis that independently evaluates the effect of curcumin supplementation on BW, WC, and BMI in diabetic patients. No evidence of publication bias was seen in this meta-analysis, as examined by the Egger and Begg regression test; in our systematic review, curcumin dose ranged from low to high; therefore, the effects of dose were considered in this systematic review. Also, the included studies were of moderate and high methodological quality. They were done on participants with the same disease. Besides, based on RAB1 quality assessment guidelines, most studies were of moderate and high quality. In addition, we could find sources of between-study heterogeneity in our subgroup analyses.

## Conclusion

In our meta-analysis, sufficient evidence was found regarding curcumin’s significant effect on BW, BMI, and WC. However, more evident doses higher than 1000 mg/day are required. Therefore, additional high-quality, well-designed studies should be performed to approve our findings.

## Declaration

## Data availability statements

All data obtained from this research are included in the article’s main text.

## Author contributions

NAL, SF, and EA conceived the study. NAL carried out the literature search and wrote the manuscript. NAL and SAL carried out data extraction and independent reviewing. NAL, EA, and SF assessed the quality of the included studies. NAL and SF performed data analysis and interpretation, and SJM revised the manuscript. The manuscript has been read and approved by all authors.

## Ethical approval and consent to participate

Not applicable.

## Consent to participate

Not applicable.

## Consent to publication

The author fully consents to the publication of the article.

